# Prenatal ochratoxin A exposure, birth outcomes and infant growth in rural Burkina Faso: a human biomonitoring sub-study from the MISAME-III trial

**DOI:** 10.1101/2024.02.23.24303256

**Authors:** Yuri Bastos-Moreira, Alemayehu Argaw, Trenton Dailey-Chwalibóg, Jasmin El-Hafi, Lionel Olivier Ouédraogo, Laeticia Celine Toe, Sarah De Saeger, Carl Lachat, Marthe De Boevre

**Affiliations:** Center of Excellence in Mycotoxicology and Public Health, MYTOX-SOUTH® Coordination Unit, Faculty of Pharmaceutical Sciences, Ghent University, Ghent, Belgium; Department of Food Technology, Safety and Health, Faculty of Bioscience Engineering, Ghent University, Ghent, Belgium; Institute of Food Chemistry, University of Münster, Münster, Germany; Laboratoire de Biologie Clinique, Centre Muraz, Bobo-Dioulasso, Burkina Faso; Unité Nutrition et Maladies Métaboliques, Institut de Recherche en Sciences de la Santé (IRSS), Bobo-Dioulasso, Burkina Faso; Department of Biotechnology and Food Technology, Faculty of Science, University of Johannesburg, Doornfontein Campus, Gauteng, South Africa

**Keywords:** birth outcomes, exposomics, growth, low- and middle-income countries, MISAME-III, mycotoxins, ochratoxin A

## Abstract

Mycotoxin exposure during pregnancy has been associated with adverse birth outcomes and poor infant growth in low- and middle-income countries. We assessed multiple biomarkers and metabolites of exposure to mycotoxins during pregnancy and their associations with birth outcomes and infant growth in 305 pregnant participants, between 30 and 34 completed weeks of gestation, in rural Burkina Faso. In this study, whole blood microsamples were analyzed for mycotoxin concentrations using ultra-performance liquid chromatography coupled to tandem mass spectrometry. Unadjusted and adjusted associations between mycotoxin exposure, and birth outcomes and infant growth at 6 months were estimated using linear regression models for continuous outcomes and linear probability models with robust variance estimation for binary outcomes. Infant growth trajectories from birth to 6 months were compared by exposure status using mixed-effects models with random intercept for the individual infant and random slope for the infant’s age. Ochratoxin A (OTA) exposure was detected in 50.8% of the study participants, with aflatoxin G1, aflatoxin M1, cyclopiazonic acid, deoxynivalenol and T-2-toxin being detected in the range of 0.33% and 2.31% of the population. We found no statistically significant (all *p* ≥ 0.05) associations between OTA exposure, and birth outcomes and infant growth. Despite this, the findings indicate a significant presence of ochratoxin A among pregnant participants. Public health policies and nutrition-sensitive interventions must ensure that OTA exposure is reduced in Burkina Faso.

## Background

Mycotoxins are secondary metabolites from fungi that grow on crops preharvest, as well as in postharvest, storage and/or in food processing (Andrews-Trevino et al., 2022; De Boevre et al., 2012; Marin et al., 2013; Pitt et al., 2000). Mycotoxin contamination commonly affects popular food items in West Africa, including chilies, maize, peanuts, and spices (Abiala et al., 2011; Ngum et al., 2022). The climate conditions of these countries, where there is an elevated temperature and humidity level, are favorable for their production (Aasa et al., 2022; Compaore et al., 2021). In low- and middle-income countries (LMICs), pregnancies frequently result in adverse outcomes such as low birth weight (LBW), preterm birth (PTB), and/or small-for-gestational age (SGA) (Lee et al., 2013). While the causes of these outcomes are multifaceted, maternal nutrition significantly impacts both pregnancy development and newborn well-being (Ota et al., 2014).

According to the International Agency for Research on Cancer (IARC), aflatoxin B1 (AFB1) is classified as carcinogenic to humans (Group 1), while fumonisin B1 (FB1) and ochratoxin A (OTA) are considered possible human carcinogens (Group 2B). Deoxynivalenol (DON) is placed in Group 3, indicating that it has not been classified regarding its carcinogenicity due to insufficient data (IARC (International Agency for Research on Cancer), 1993). It has been suggested that aflatoxins (AFs) and DON hinders protein synthesis resulting in a modified intestinal structure, while fumonisins (FBs) reduce complex sphingolipids. Formerly, a high AFs exposure in newborns has resulted in stunting and/or underweight, while those with a high FBs exposure were also shorter and lighter (Lombard, 2014). In addition, research has also shown that DON can cross the placental barrier in humans (Nielsen et al., 2011; Tan et al., 2023), and is consequently associated with decreased birth weight (Coronel et al., 2010).

OTA has a half-life of 35.6 days and is reported to have toxic effects in humans including carcinogenicity, immunotoxicity and nephrotoxicity (Coronel et al., 2010). OTA serves as an effective biomarker of exposure owing to its persistence in blood, as it swiftly binds to plasma proteins with high affinity (Coronel et al., 2010). Previous studies have also revealed transplacental transfer of mycotoxins such as OTA to the fetus in humans (Woo et al., 2012). This exposure during the critical first 1,000 days of life (Groopman et al., 2014) might contribute to suboptimal fetal and infant outcomes (Etzel & Zilber, 2014). Nevertheless, the evidence for the association between mycotoxin exposure during pregnancy and newborn and infant growth outcomes remains inconsistent and are mainly limited with AFs and FBs. While certain studies have documented an association between exposure to certain mycotoxin groups and a higher risk of adverse birth outcomes and suboptimal infant growth, others have indicated no clear association or have even suggested a positive correlation with improved growth (Gönenç et al., 2020; Kyei et al., 2020; Tesfamariam et al., 2020).

The presence of mycotoxins in sub-Saharan populations is mainly due to monotonous diets based on contaminated staple food crops (Bankole & Adebanjo, 2003). In Burkina Faso, cereals are consumed in large quantities and have a social, economic, and nutritional significance. Maize is the second most cereal produced, with production reaching 1,710,898 tons in 2019 (Somda et al., 2023). However, in Burkina Faso, the availability of biological and toxicological data on food contamination remains limited (Kpoda et al., 2022), and the implementation of legislation and regulations concerning mycotoxins is lacking (FAO, 2003; Warth et al., 2012). Using data from the Biospecimen (BioSpé) sub-study of the MISAME-III (MIcronutriments pour la SAnté de la Mère et de l’Enfant) trial in rural Burkina Faso, we aimed to quantify maternal mycotoxin exposure during pregnancy and investigated the association with birth outcomes and infant growth.

## Methods

### Study setting, participants, and design

Study protocols for the main MISAME-III trial (Vanslambrouck et al., 2021) and the BioSpé sub-study nested under the MISAME-III trial (Bastos-Moreira et al., 2023) were published previously. The research took place within the catchment areas of six rural health centers located in the Houndé district of Burkina Faso. These areas have a Sudano-Sahelian climate, marked by a dry season lasting from September/October to April, with farming serving as the primary source of income.

The objective of the BioSpé research was to examine the impact of a multi-micronutrient enriched BEP supplement on pregnant individuals and their infants, aiming to assess the physiological effects of the supplementation on maternal health, infant birth and growth outcomes. This evaluation was conducted through comprehensive multi-omics analyses, human biomonitoring of contaminants and assessment of biological aging markers (Bastos-Moreira et al., 2023; Hanley-Cook et al., 2023). In the present study, we report the associations between maternal exposure to mycotoxins during pregnancy and various growth and nutritional status outcomes at birth and during infancy.

### Exposure and outcomes

The present study considered exposure to a range of mycotoxins listed in Table 2. However, aflatoxin B1 (AFB1)-lysine exposure was not assessed due to the current unavailability of the commercial analytical standard for this specific mycotoxin. Mycotoxin exposure was determined as the presence of a concentration equal to or greater than the limit of detection (LOD) in whole blood microsamples.

The outcomes of interest were birth outcomes, such as birth weight, SGA, LBW, gestational age (GA), PTB, length, mid-upper arm circumference (MUAC), head circumference, ponderal index, chest circumference, and infant growth and nutritional status at the age of 6 months, such as LAZ, weight-for-age (WAZ) and weight-for-length *z*-score (WLZ), MUAC, head circumference, hemoglobin, stunting, underweight wasting and anemia. We additionally assessed the associations between mycotoxin exposure and infant growth trajectories (height, weight, upper-arm and head circumferences) during the first 6 months postpartum.

PTB was characterized as the delivery of a newborn before completing 37 weeks of gestation. SGA was defined as a newborn weight less than the 10^th^ percentile of weight for the same GA and sex, as per the International Fetal and Newborn Growth Consortium for the 21^st^ Century (INTERGROWTH-21^st^) (Villar et al., 2014). Anthropometric *z*-scores of LAZ, WAZ and WLZ were calculated based on the WHO Child Growth Standards with stunting, underweight and wasting defined as LAZ, WAZ and WLZ values below 2 SD from the median value for same age and sex from the reference population (de Onis & Branca, 2016). Newborn Rohrer’s ponderal index was calculated as weight in g divided for length in cm cubed (i.e., weight/length^3^ (g/cm^3^) × 1,000).

### Data collection

The MISAME-III trial data were collected through computer-assisted personal interviewing using SurveySolutions (version 21.5) on tablets and then transferred to a central server at Ghent University. Sociodemographic and other relevant characteristics of participants and study households were collected at baseline during the first and third trimester of pregnancy. All newborn anthropometry measurements were taken within 12 hours of birth, whereas mothers were invited for follow-up growth assessment every month until 6 months of age. Measurements were conducted in duplicates, with a third measurement taken if a significant discrepancy was observed between the duplicate measurements. Length was assessed using a Seca 416 Infantometer, measuring to the nearest 1 mm, while weight was measured with a Seca 384 scale, accurate to the nearest 10 g. Head circumference, thoracic circumference, and MUAC were measured to the nearest 1 mm using a Seca 212 measuring tape. GA was determined utilizing a portable ultrasound (SonoSite M-Turbo, FUJIFILM SonoSite, Bothell, Washington, USA) during the first and early stages of the second trimester of pregnancy.

### Biospecimen collection through volumetric absorptive microsampling

The samples collection procedure was described in detail previously (Bastos-Moreira et al., 2023). Pregnant participants (n=305) were invited to their local health center at 30-34 weeks of gestation, between March and July 2021. Since volumetric absorptive microsampling (VAMS) collection for the different analyses in the study were conducted in the same visit, to preserve metabolites for metabolomics, participants were advised to fast on the morning of VAMS collection. A total amount of 40 µL (2 × 20 µL VAMS tips) of capillary whole blood was collected by capillary sampling onto a VAMS tip, namely Mitra^TM^, via direct finger incision for mycotoxins analysis.

Then, 20 µL Mitra Clamshells were transported from the health centers to the Institut de Recherche en Sciences de la Santé in Bobo-Dioulasso, Burkina Faso for shipment at room temperature to Ghent University, Belgium. For storage at −80°C until analysis, VAMS were placed in Mitra Autoracks inside a storage bag containing desiccant bags.

Items used for VAMS collection were purchased from Neoteryx (Torrance, CA, USA).

### Sample extraction procedure

#### Preparation of calibration curve

For the method development, ethylenediaminetetraacetic acid anticoagulated blood were supplied by Red Cross Flanders (Ghent, Belgium) and kept at −80 °C until use. These blood samples were pipetted in 2 mL Eppendorf tubes and fortified (spiked) with a mix of 35 mycotoxins at five different concentration levels. Then, each calibration curve point was prepared by dipping a VAMS tip into the designated pool of spiked whole blood concentration. After the tip was fully colored, contact with the whole blood was maintained for 7 seconds to obtain full absorption. Overfilling of the VAMS devices was avoided by not completely immersing the tip into the whole blood, as described previously (Vidal et al., 2021). The devices were subsequently dried in their accompanying Mitra^TM^ clamshells for ≥3 hours at room temperature.

#### Mycotoxins extractions from VAMS

A VAMS multi-mycotoxin extraction methodology was previously established in our laboratory (Vidal et al., 2021). Briefly, extraction began by transferring the VAMS tips from the plastic handles into 2 mL Eppendorf tubes, and pipetting 250 μL extraction solvent (acetonitrile/water/acetic acid, 59/40/1, *v*/*v*/*v*), containing the internal standards 0.125 µg/L ^13^C_17_–AFB1 and 0.25 µg/L for ^13^C_15_ – deoxynivalenol (DON), ^13^C_34_–FB1 and 0.125 µg/L for ^13^C_18_–zearalenone, to the sample tubes. Subsequently, samples were ultrasonicated for 30 minutes and shaken for 60 minutes at 25°C with rotation at 1,400 rpm in a Biosan TS-100 Thermo-Shaker followed by centrifugation (10 minutes at 10,000g, room temperature). The tips were discarded, and the supernatant was pipetted to an 8 mL glass tube and evaporated under nitrogen on a Turbovap LV Evaporator (Biotage, Charlotte, USA).

Afterwards, the extracts were reconstituted in 50 μL of injection solvent (methanol/water, 60/40, *v*/*v*), vortexed, centrifuged (for 10 min at 5000 g) and filtered (22 μm, PVDF, Durapore^®^,Cork, Ireland). Lastly, samples were transferred into vials before 10 μL were injected into the ultra-performance liquid chromatography coupled to tandem mass spectrometry (UPLC -MS/MS) instrument.

#### UPLC-MS/MS analysis

Sample analysis was conducted using a Waters UPLC^®^ system coupled to a Quattro XEVO TQ-XS mass spectrometer (Waters, Manchester, UK). Detailed instrument parameters can be found in a previous study (Vidal et al., 2021). Data acquisition and processing were carried out using MassLynx™ version 4.1 and QuanLynx^®^ version 4.1 software (Waters, Manchester, UK).

#### Validation of the VAMS-based UPLC-MS/MS method

The UPLC-MS/MS method validation experiment was conducted in three independent replicates over 4 different days. The method validation was re-validated from a previous method (Vidal et al., 2021) to optimize the extraction protocol and mass spectrometry parameters prior to analysis of BioSpé samples. The method was validated following the guidelines of the European Commission (EC) decision No. 2002/657 (European Commission, 2002) and Commission Regulation (EU) No. 401/2006 (European Commission, 2006).

Whole blood samples supplied by Red Cross Flanders were spiked at five different concentration levels to determine the limit of detection (LOD) and lower limit of quantification (LLOQ). LLOQ is the concentration at which the analytical method cannot operate with acceptable precision (Currie, 1995; Thompson et al., 2002). Apparent recoveries were attained by comparing the detected concentrations (calculated using the analyzed calibration curve in pure solvent) against the spiked concentrations. The mean and standard deviation of the detected concentrations were calculated for each concentration level. The intraday precision (repeatability (RSDr)) and interday precision intra-laboratory (reproducibility (RSDR) of the method were expressed as the variation coefficient (CV %).

#### Statistical analysis

Data cleaning and statistical analyses were conducted using Stata (Stata Statistical Software: release 17; StataCorp), with statistical significance evaluated at a two-sided significance level of *p* < 0.05. Descriptive statistics are presented using means ± SD or medians (range) for the continuous variables, depending on the nature of the data distribution, and frequencies (percentages) for nominal variables.

In the study sample, only exposure to OTA was found in an adequate number of participants to assess the association with newborn and infant outcomes. We assessed the association between OTA exposure and the study outcomes at birth and 6 months of age and employed linear regression models to analyze continuous outcomes and linear probability models with robust variance estimation for binary outcomes. All models were adjusted for clustering by the health center catchment areas and allocation for the prenatal and postnatal BEP interventions. Furthermore, adjusted models additionally included the covariates maternal age, primiparity, baseline BMI and hemoglobin concentration, and household size, wealth index score, access to improved water and sanitation, and food security status.

We also compared OTA exposed and non-exposed groups by growth trajectories from birth to 6 months. For this purpose, We employed mixed-effects regression models with a random intercept for each individual infant and a random slope for the infant’s age in months. We investigated the optimal growth trajectory that aligns with our data by visually examining graphs and comparing model fit indices, such as AIC (Akaike Information Criterion) and BIC (Bayesian Information Criterion) values. As a result, we utilized quadratic models for height, weight, and MUAC, along with a restricted cubic spline model with 4 knots for head circumference. We accounted for the correlation among repeated measurements within an individual by considering an unstructured covariance matrix. Fixed effects in the model included the main effect of exposure status, age, and exposure by age interaction, estimating the difference in monthly changes in the outcome between exposure and unexposed groups. Models were further adjusted for the aforementioned covariates.

## Results

### Participants characteristics

Of the 305 study participants, birth outcomes and infant growth at 6 months were assessed on 296 and 274 participants, respectively (**Figure 1**). Mean ± SD age of the study participants was 24.1 ± 5.61 years and 45.6% of the mothers had at least a primary education (**Table 1**). The mean ± SD maternal BMI at study inclusion was 22.0 ± 3.17 kg/m^2^ with 7.77% underweight (BMI<18.5 kg/m^2^). More than two-thirds (70.1%) of the mothers were from food insecure households, 32.1% were anemic, and the mean ± SD maternal dietary diversity score during pregnancy was 4.05 ± 1.29 food groups.

**Table 1.**
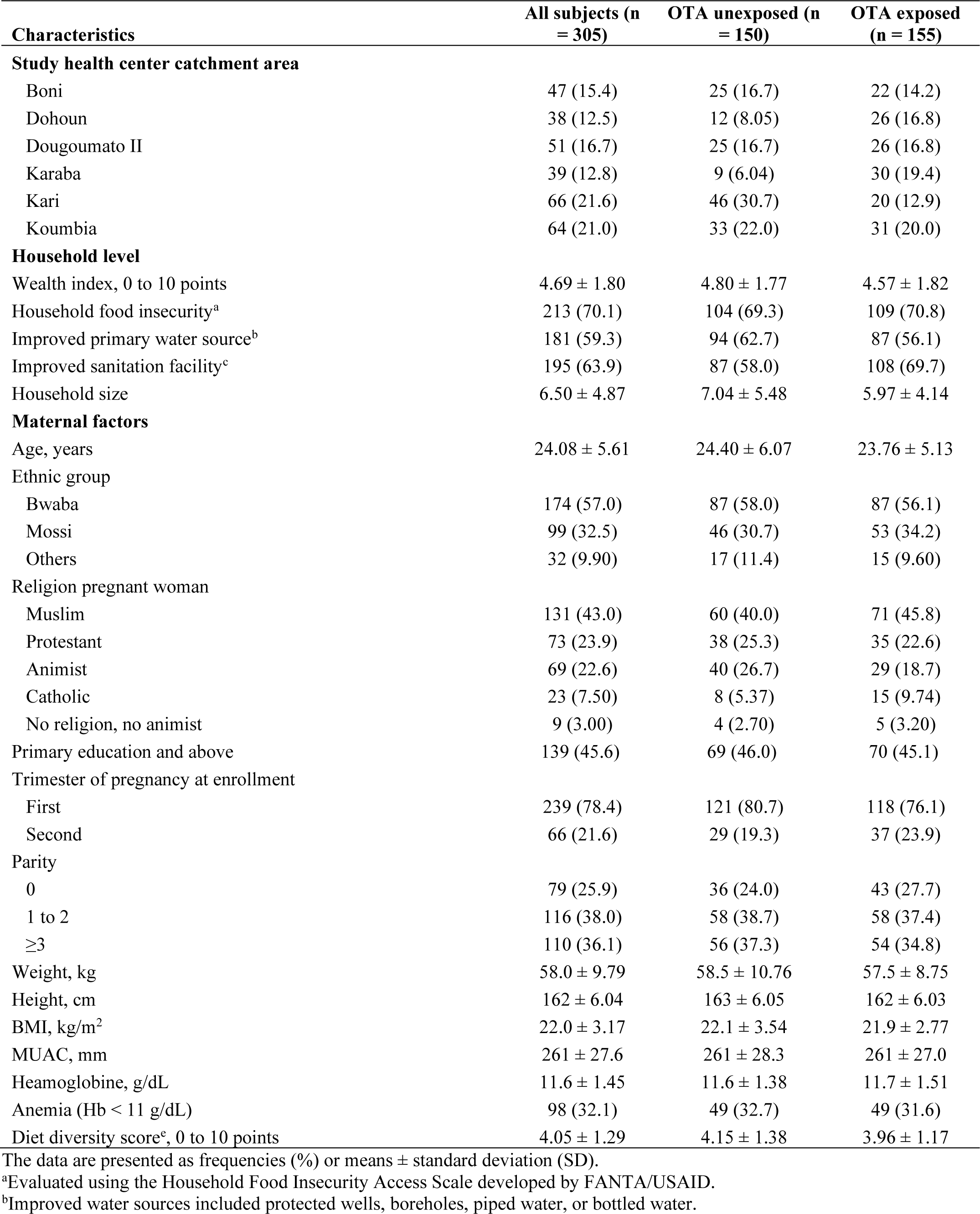
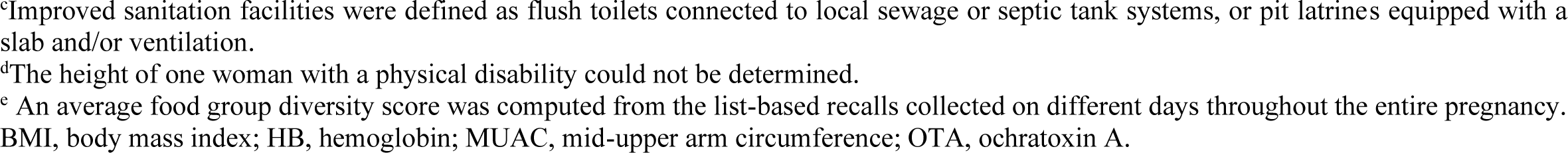
Characteristics of study participants.

| Characteristics | All subjects (n<br>= 305) | OTA unexposed (n<br>= 150) | OTA exposed<br>(n = 155) |
| --- | --- | --- | --- |
| **Study health center catchment area** |  |  |  |
| Boni | 47 (15.4) | 25 (16.7) | 22 (14.2) |
| Dohoun | 38 (12.5) | 12 (8.05) | 26 (16.8) |
| Dougoumato II | 51 (16.7) | 25 (16.7) | 26 (16.8) |
| Karaba | 39 (12.8) | 9 (6.04) | 30 (19.4) |
| Kari | 66 (21.6) | 46 (30.7) | 20 (12.9) |
| Koumbia | 64 (21.0) | 33 (22.0) | 31 (20.0) |
| **Household level** |  |  |  |
| Wealth index, 0 to 10 points | 4.69 ± 1.80 | 4.80 ± 1.77 | 4.57 ± 1.82 |
| Household food insecurity<sup>a</sup> | 213 (70.1) | 104 (69.3) | 109 (70.8) |
| Improved primary water source<sup>b</sup> | 181 (59.3) | 94 (62.7) | 87 (56.1) |
| Improved sanitation facility<sup>c</sup> | 195 (63.9) | 87 (58.0) | 108 (69.7) |
| Household size | 6.50 ± 4.87 | 7.04 ± 5.48 | 5.97 ± 4.14 |
| **Maternal factors** |  |  |  |
| Age, years | 24.08 ± 5.61 | 24.40 ± 6.07 | 23.76 ± 5.13 |
| Ethnic group |  |  |  |
| Bwaba | 174 (57.0) | 87 (58.0) | 87 (56.1) |
| Mossi | 99 (32.5) | 46 (30.7) | 53 (34.2) |
| Others | 32 (9.90) | 17 (11.4) | 15 (9.60) |
| Religion pregnant woman |  |  |  |
| Muslim | 131 (43.0) | 60 (40.0) | 71 (45.8) |
| Protestant | 73 (23.9) | 38 (25.3) | 35 (22.6) |
| Animist | 69 (22.6) | 40 (26.7) | 29 (18.7) |
| Catholic | 23 (7.50) | 8 (5.37) | 15 (9.74) |
| No religion, no animist | 9 (3.00) | 4 (2.70) | 5 (3.20) |
| Primary education and above | 139 (45.6) | 69 (46.0) | 70 (45.1) |
| Trimester of pregnancy at enrollment |  |  |  |
| First | 239 (78.4) | 121 (80.7) | 118 (76.1) |
| Second | 66 (21.6) | 29 (19.3) | 37 (23.9) |
| Parity |  |  |  |
| 0 | 79 (25.9) | 36 (24.0) | 43 (27.7) |
| 1 to 2 | 116 (38.0) | 58 (38.7) | 58 (37.4) |
| ≥3 | 110 (36.1) | 56 (37.3) | 54 (34.8) |
| Weight, kg | 58.0 ± 9.79 | 58.5 ± 10.76 | 57.5 ± 8.75 |
| Height, cm | 162 ± 6.04 | 163 ± 6.05 | 162 ± 6.03 |
| BMI, kg/m<sup>2</sup> | 22.0 ± 3.17 | 22.1 ± 3.54 | 21.9 ± 2.77 |
| MUAC, mm | 261 ± 27.6 | 261 ± 28.3 | 261 ± 27.0 |
| Heamoglobine, g/dL | 11.6 ± 1.45 | 11.6 ± 1.38 | 11.7 ± 1.51 |
| Anemia (Hb < 11 g/dL) | 98 (32.1) | 49 (32.7) | 49 (31.6) |
| Diet diversity score<sup>e</sup>, 0 to 10 points | 4.05 ± 1.29 | 4.15 ± 1.38 | 3.96 ± 1.17 |

The data are presented as frequencies (%) or means ± standard deviation (SD).
<sup>a</sup>Evaluated using the Household Food Insecurity Access Scale developed by FANTA/USAID.
<sup>b</sup>Improved water sources included protected wells, boreholes, piped water, or bottled water.
<sup>c</sup>Improved sanitation facilities were defined as flush toilets connected to local sewage or septic tank systems, or pit latrines equipped with a slab and/or ventilation.
<sup>d</sup>The height of one woman with a physical disability could not be determined.
<sup>e</sup> An average food group diversity score was computed from the list-based recalls collected on different days throughout the entire pregnancy. BMI, body mass index; HB, hemoglobin; MUAC, mid-upper arm circumference; OTA, ochratoxin A.

**Figure 1.**
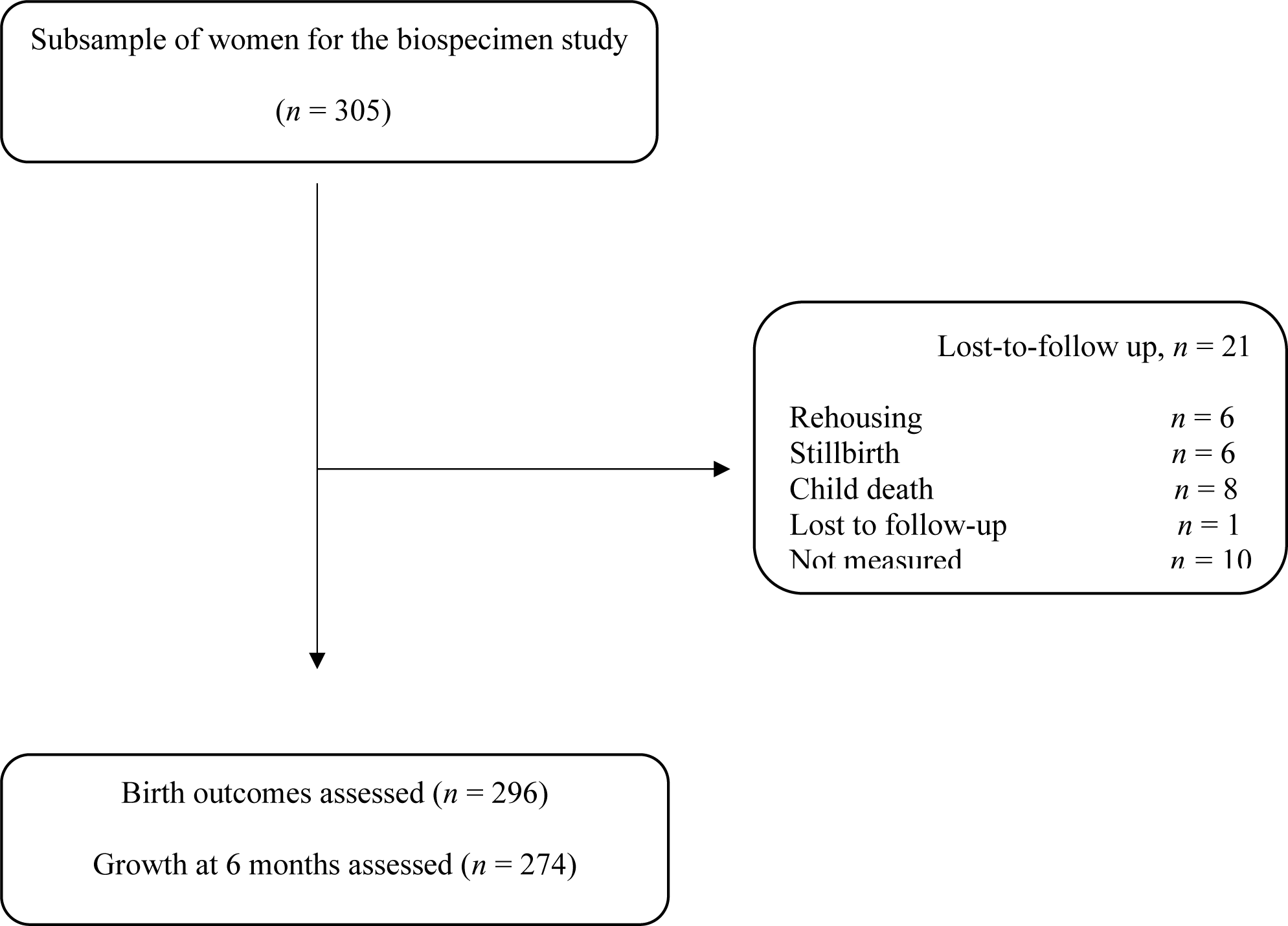
Study flow diagram of the Biospecimen sub-study (BioSpé) of the MISAME-III project.

### Mycotoxins exposure and newborn and infant growth and nutritional status

The microsample (VAMS sample) whole blood analysis using the ultra-performance liquid-performance chromatography-mass spectrometry analysis showed that, aside from OTA, almost all participants were found to be not exposed to most mycotoxins (**Table 2**). OTA exposure was detected in 50.8% of the study participants with a median (range) concentration of 0.01 (<LOD, 8.21) µg/L. Other mycotoxins including aflatoxin G (AFG1), aflatoxin M1 (AFM1), cyclopiazonic acid (CPA), DON and T-2-toxin were detected in the range of 2.31% and 0.33% of the population. For the remaining mycotoxins, no exposure was detected through whole blood analysis.

**Table 2.** Mycotoxin exposure during the third trimester of pregnancy.

| **Mycotoxins** | **Positive samples: n (%)** | **Median (range) µg/L** |
| --- | --- | --- |
| 15-acetyldeoxynivalenol | 0.00 | <LOD (<LOD, <LOD) |
| 3-acetyldeoxynivalenol | 0.00 | <LOD (<LOD, <LOD) |
| aflatoxin B1 | 0.00 | <LOD (<LOD, <LOD) |
| aflatoxin B2 | 0.00 | <LOD (<LOD, <LOD) |
| aflatoxin G1 | 7 (2.31) | <LOD (<LOD, 0.22) |
| aflatoxin G2 | 0.00 | <LOD (<LOD, <LOD) |
| aflatoxin M1 | 2 (0.66) | <LOD (<LOD, 1.61) |
| alpha-zearalenol | 0.00 | <LOD (<LOD, <LOD) |
| alternariol | 0.00 | <LOD (<LOD, <LOD) |
| alternariol monomethyl ether | 0.00 | <LOD (<LOD, <LOD) |
| beauvericin | 0.00 | <LOD (<LOD, <LOD) |
| beta- zearalenol | 0.00 | <LOD (<LOD, <LOD) |
| citrinin | 0.00 | <LOD (<LOD, <LOD) |
| cyclopiazonic acid | 4 (1.32) | <LOD (<LOD, 2.77) |
| deepoxy- deoxynivalenol | 0.00 | <LOD (<LOD, <LOD) |
| deoxynivalenol -3- glucoside | 0.00 | <LOD (<LOD, <LOD) |
| deoxynivalenol | 1 (0.33) | <LOD (0.00, 0.60) |
| diacetoxyscirpenol | 0.00 | <LOD (<LOD, <LOD) |
| enniatin A | 0.00 | <LOD (<LOD, <LOD) |
| enniatin A1 | 0.00 | <LOD (<LOD, <LOD) |
| enniatin B | 0.00 | <LOD (<LOD, <LOD) |
| enniatin B1 | 0.00 | <LOD (<LOD, <LOD) |
| fumonisin B1 | 0.00 | <LOD (<LOD, <LOD) |
| fumonisin B2 | 0.00 | <LOD (<LOD, <LOD) |
| fumonisin B3 | 0.00 | <LOD (<LOD, <LOD) |
| fusarenone-X | 0.00 | <LOD (<LOD, <LOD) |
| neosolaniol | 0.00 | <LOD (<LOD, <LOD) |
| nivalenol | 0.00 | <LOD (<LOD, <LOD) |
| ochratoxin A | 154 (50.83) | 0.01 (<LOD, 8.21) |
| ochratoxin alpha | 0.00 | <LOD (<LOD, <LOD) |
| roquefortine-C | 0.00 | <LOD (<LOD, <LOD) |
| sterigmatocystine | 0.00 | <LOD (<LOD, <LOD) |
| T-2-toxin | 4 (1.32) | <LOD (<LOD, 0.75) |
| zearalanone | 0.00 | <LOD (<LOD, <LOD) |
| zearalenone | 0.00 | <LOD (<LOD, <LOD) |

LOD, limit of detection

There were no statistically significant associations between maternal OTA exposure during pregnancy and the duration of gestation or size at birth (**Table 3**). Similarly, OTA exposure was not significantly associated with the aforementioned infant growth and nutritional status at the age of 6 months (**Table 4**). We also found no significant association between OTA exposure and growth trajectories from birth to the age of 6 months (**Figure 2**).

**Table 3.** Maternal ochratoxin A exposure during pregnancy and birth outcomes^1^.

| Outcomes | OTA unexposed (n = 146) | OTA exposed (n = 150) | Unadjusted beta (95% CI) | *p* | Adjusted beta (95% CI) | *p* |
| --- | --- | --- | --- | --- | --- | --- |
| Birthweight, kg | 3.00 ± 0.50 | 3.01 ± 0.42 | 0.01 (-0.11, 0.12) | 0.909 | 0.15 (-0.09, 0.12) | 0.779 |
| Low birthweight (<2.5 kg) | 20 (13.7) | 20 (13.3) | -0.28 (-8.18, 7.62) | 0.944 | -0.06 (-7.64, 7.53) | 0.988 |
| Small-for-gestational age | 32 (21.9) | 45 (30.0) | 6.94 (-3.27, 17.2) | 0.182 | 6.62 (-3.52, 16.7) | 0.200 |
| Gestational age, week | 39.8 ± 1.68 | 39.9 ± 1.39 | 0.19 (-0.18, 0.56) | 0.325 | 0.19 (-0.18, 0.56) | 0.309 |
| Preterm delivery (<37 week) | 8 (5.48) | 4 (2.61) | -2.68 (-7.25, 1.89) | 0.249 | -2.53 (-7.22, 2.16) | 0.289 |
| Length, cm | 48.3 ± 2.40 | 48.6 ± 1.96 | 0.25 (-0.25, 0.76) | 0.326 | 0.29 (-0.20, 0.79) | 0.243 |
| MUAC, mm | 99.6 ± 8.95 | 100.5 ± 8.60 | 1.08 (-0.90, 3.07) | 0.283 | 1.02 (-0.84, 2.88) | 0.280 |
| Head circumference, cm | 33.2 ± 1.66 | 33.4 ± 1.38 | 0.26 (-0.09, 0.61) | 0.144 | 0.28 (-0.06, 0.62) | 0.102 |
| Ponderal index, gm/cm<sup>3</sup> | 26.4 ± 2.57 | 26.1 ± 2.42 | -0.28 (-0.85, 0.29) | 0.333 | -0.28 (-0.83, 0.26) | 0.308 |
| Chest circumference, cm | 31.7 ± 2.00 | 31.9 ± 1.64 | 0.18 (-0.26, 0.62) | 0.427 | 0.20 (-0.21, 0.61) | 0.335 |

<sup>1</sup>Values are mean ± SD or frequencies (percentages). Beta coefficients, both unadjusted and adjusted, are computed using linear regression models to analyze continuous outcomes and linear probability models with robust variance estimation for the binary outcomes. Every model was adjusted for the health center catchment areas and the prenatal and postnatal interventions arms, while adjusted models additionally contained maternal age, primiparity, baseline BMI and hemoglobin concentration, and household size, wealth index score, access to improved water and sanitation, and food security status. MUAC, mid-upper arm circumference; OTA, ochratoxin A.

**Table 4.**
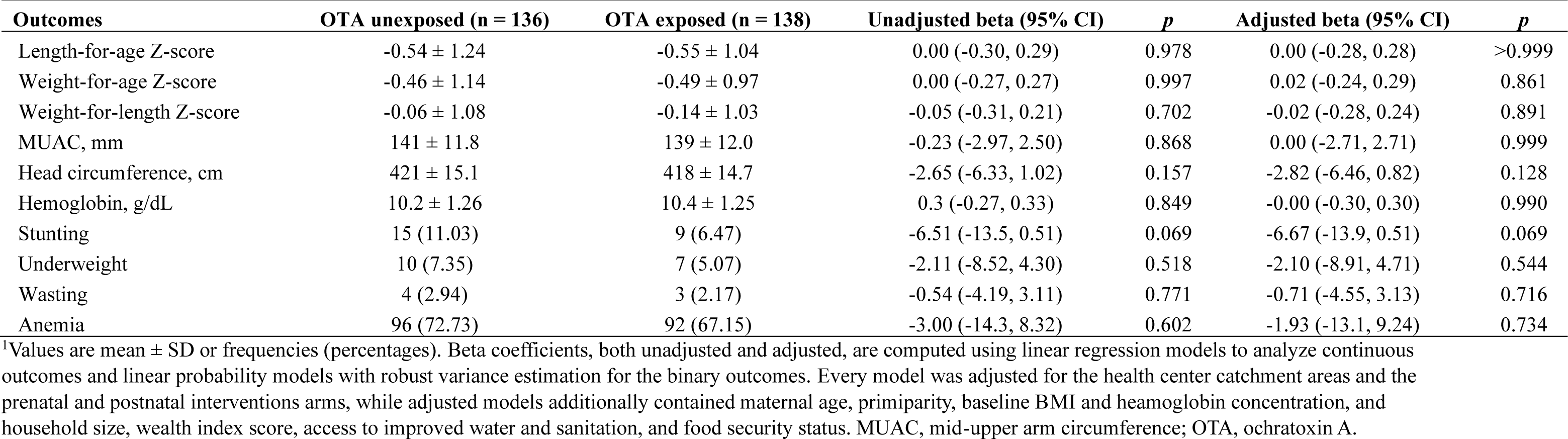
Maternal ochratoxin A exposure and infant growth and nutritional status at 6 months of age^1^.

**Figure 2.**
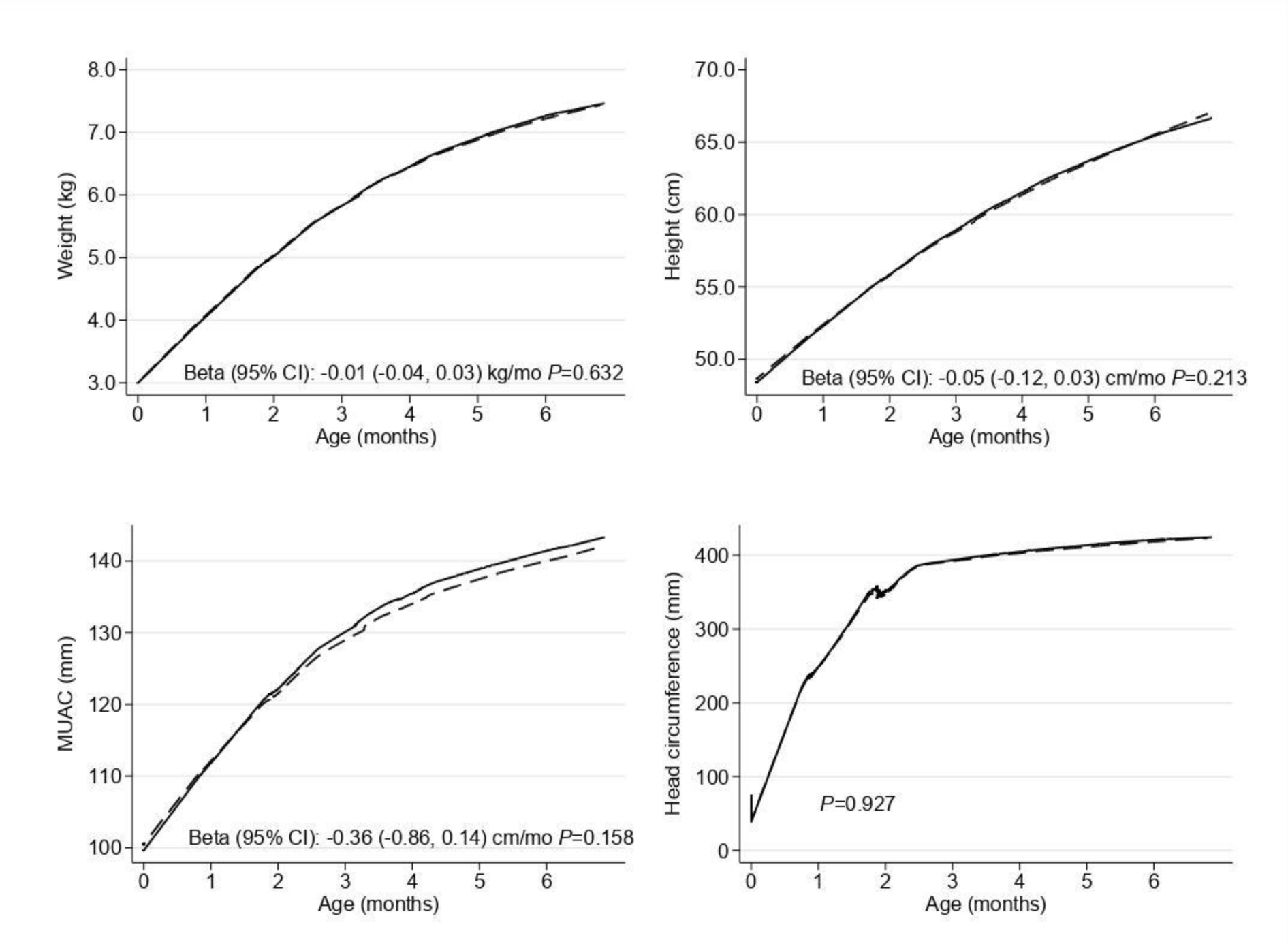
Infant growth trajectories from birth to 6 months by ochratoxin A unexposed groups (solid lines) and exposed (dashed lines)

### Validity of the UPLC-MS/MS

The described UPLC-MS/MS conditions were sufficient for OTA determination (Figure 3). Calibration curves were established with correlation coefficient >0.95. The LODs of the various mycotoxins metabolites/biomarkers ranged in the low µg/L level from 0.05 (AFG1), 0.11 (AFM1), 1.44 (CPA), 0.40 (DON), 0,09 (OTA), to 0.59 µg/L (T-2-toxin). The LLOQs were 0.09 (AFG1), 0.20 (AFM1), 2.65 (CPA), 0,80 (DON), 0.17 (OTA), to 1.10 (T-2 Toxin). Satisfactory apparent recoveries were achieved for all detected mycotoxins with the mean values of 89% −105%. The acceptable RSDr and RSDR for the different mycotoxins and metabolites was <15.

**Figure 3.**
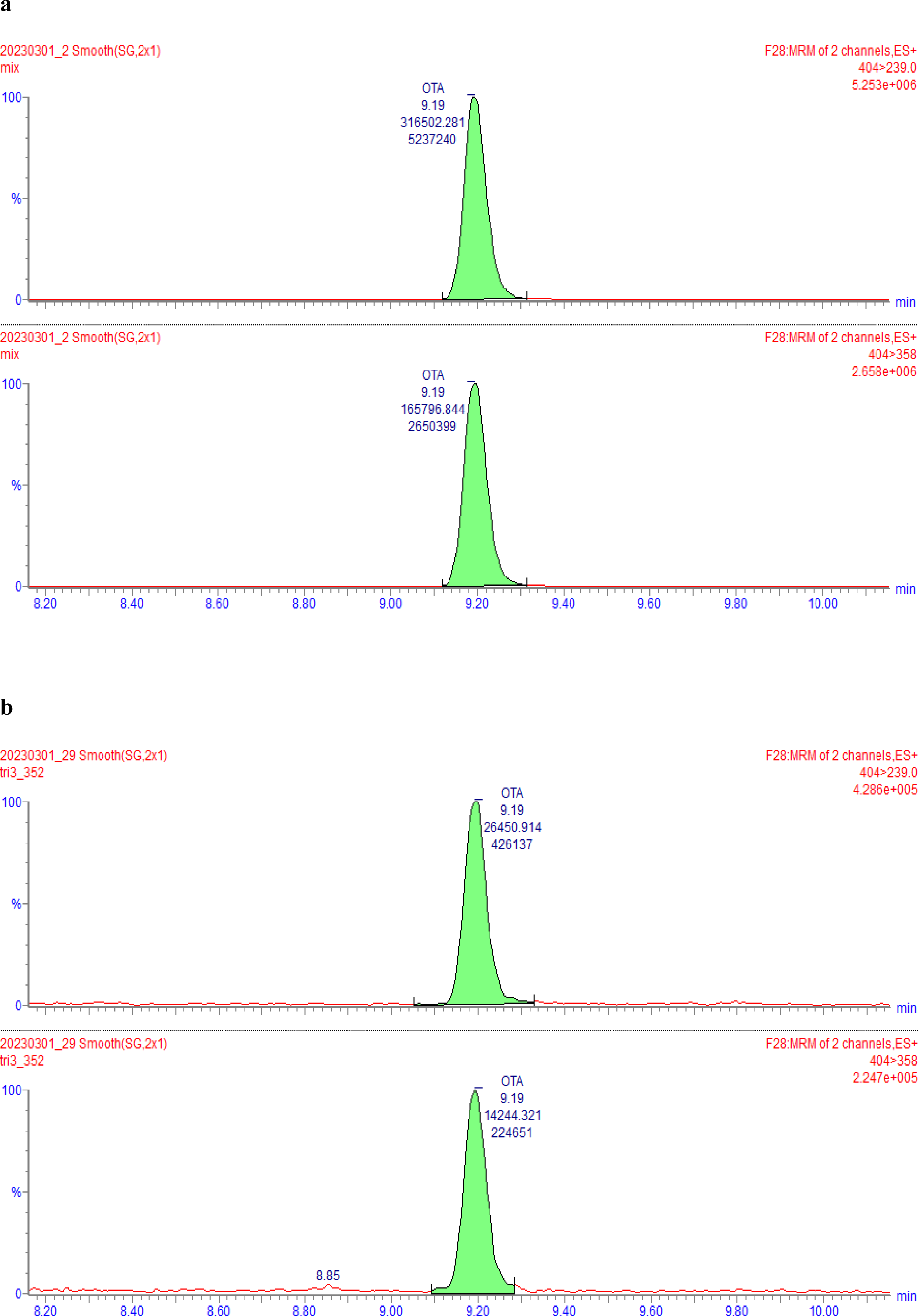
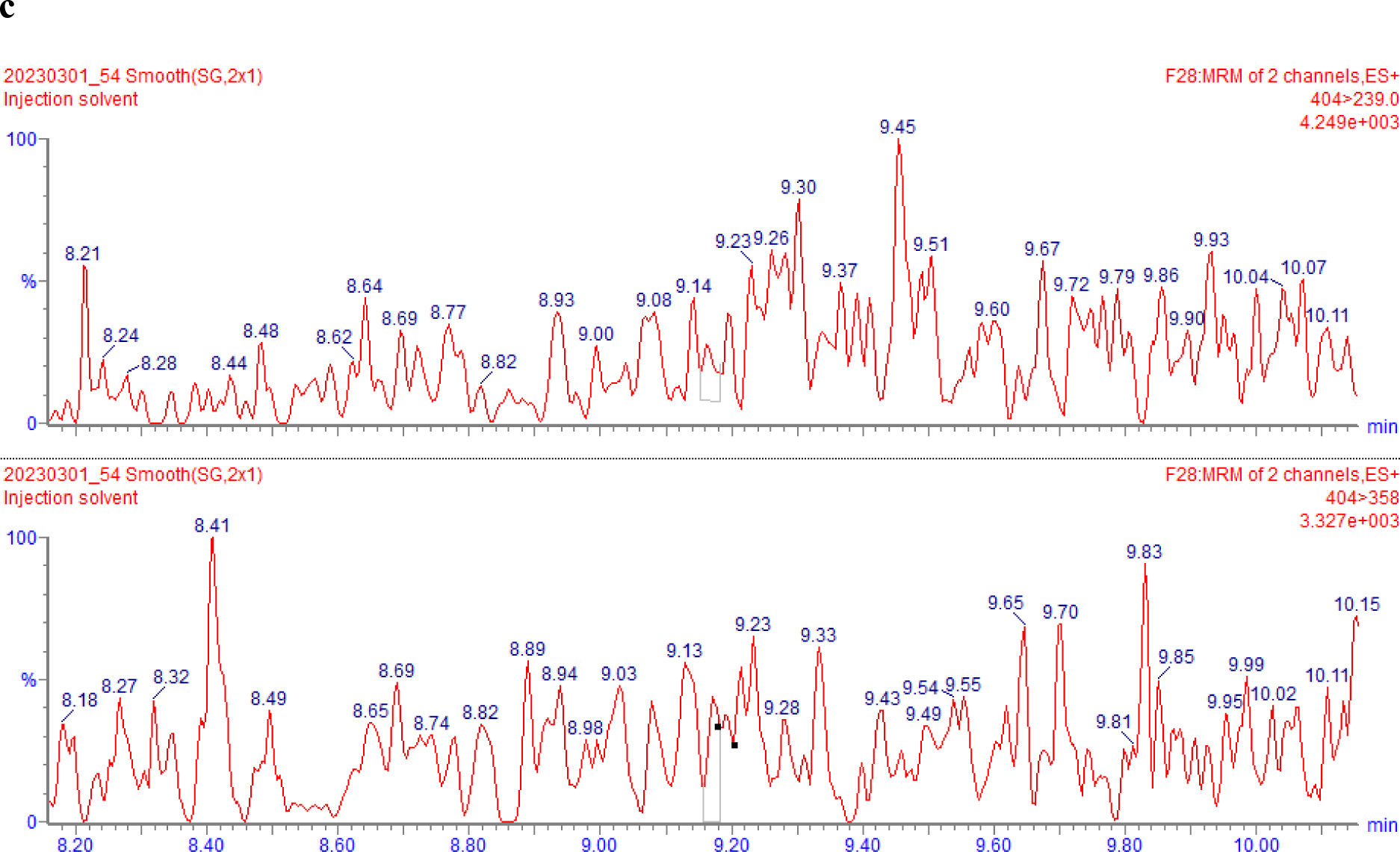
UPLC-MS/MS chromatograms of (A) OTA standard solution 2 µg/L; (B) OTA-naturally contaminated whole blood microsample (concentration 0.89 µg/L); (C) OTA-free whole blood microsample.

## Discussion

The present study found a high prevalence of OTA exposure (50.8%) during pregnancy. Exposures to mycotoxins, such as AFG1, AFM1, CPA, DON and T-2-toxin, were detected in relatively fewer subjects, whereas other mycotoxins were not detected. We found no apparent association between maternal OTA exposure during pregnancy and newborn anthropometry, and infant growth and nutritional status.

OTA is mainly produced by select species of Aspergillus, Monascus, and Penicillium, and is often found contaminating food sources (Alshannaq & Yu, 2017; Frisvad et al., 2005). As a result, avoiding dietary exposure to OTA is challenging due to the widespread occurrence of these toxins in various food stuffs (Pfohl-Leszkowicz & Manderville, 2007). In Burkina Faso, Ware *et al*. (2017) noted OTA contamination in 7.5% of infant formulas derived from maize. The highest concentration recorded was in a sample containing a mixture of maize, beans, peanuts and soya (3.2 mg/kg). In the papers reviewed by Arce-López *et al*. (2020), OTA was of the most frequently detected mycotoxins in blood, plasma and serum samples (Arce-López et al., 2020). Authors reported incidence levels of 64.9% (Ali et al., 2018; Fan et al., 2019; Karima et al., 2010; Tesfamariam et al., 2022), and concluded that the global population is generally exposed to OTA due to its long half-life in these matrices (Arce-López et al., 2020). In addition, the occurrence of OTA detected in the present study is comparable to previous literature. Fan *et al*. (2019) analyzed multi-mycotoxin exposure in plasma samples of 260 adults in China and detected OTA as the most abundant mycotoxin detected in 27.7% of the participants (range: 0.31-9.18 µg/L) (Fan et al., 2019), and in another study the OTA prevalence was 28% of 107 serum samples from Tunisia (range: 0.12 and 11.67 µg/L) (Karima et al., 2010). Likewise, in plasma samples from Bangladesh, OTA concentrations were between 0.10 and 0.87 µg/L (mean 0.35 ± 0.20 µg/L) in women, while for males, the values were 0.16–3.07 µg/L (mean 0.70 ± 0.71 µg/L) (Ali et al., 2018). The detection and quantification limits for these studies were 0.04 and 0.10 µg/L, 0.l0 and 0.20 µg/L and 0.05 and 0.10 µg/L respectively (Ali et al., 2018; Fan et al., 2019; Karima et al., 2010).

On the other hand, exposure to mycotoxins other than OTA was found to be low in the study population as compared to what has been reported previously in LMICs (Abdulrazzaq et al., 2002; Lauer et al., 2020; Mahdavi et al., 2010; Tesfamariam et al., 2022). The variations in physicochemical properties of mycotoxins lead to differences in their toxicokinetic profiles. This results in a wide range of excretion amounts and times, spanning from hours to days, and from parts per billion to parts per trillion concentrations. It is conceivable that the used UPLC-MS/MS system may not detect the lowest concentrations of mycotoxin metabolites. Additionally, participants were instructed to fast on the morning of VAMS collection, since Mitra tips were used for both metabolomics and mycotoxin analysis simultaneously. This fasting period may have accelerated the metabolism of certain mycotoxins, hence it is imperative to investigate the stability of mycotoxins during food processing, their behavior within the digestive system, and conduct toxicodynamic and toxicokinetic studies (McCormick et al., 2015). Additionally, understanding the formation process of these metabolites, along with comprehension of their structure and molecular mass, is the key to resolving the analytical and technological hurdles associated with them. Therefore, given these facts, there may be underreporting of exposure to certain mycotoxins due to their short-half lives.

No statistically significant associations were found between OTA exposure and birth outcomes, including gestational duration or birth size. Similarly, OTA exposure was not significantly associated with infant growth and nutritional status. OTA exposure has been previously reported to be unassociated to anthropometric data (Akdemir et al., 2010; Erdal & Yalçın, 2022; Jonsyn-Ellis, 2000; Silva et al., 2020) other than body weight (Skaug et al., 2001). On the other hand, an association between poor birth outcomes and infant growth and exposure to other mycotoxin groups have been reported inconsistently. For instance, a study conducted in Ethiopia discovered an association between chronic maternal exposure to AF and decreased fetal growth trajectories, as determined by fetal biometry derived from ultrasound estimates. However, the same study did not report an association of AF exposure with birth anthropometry (Tesfamariam et al., 2022). Yousef *et al*. (2002) also did not find any significant correlation between AFM1 exposure and GA, postnatal age, clinical condition or gender in the United Arab Emirates. A systematic review of studies that evaluated mycotoxin exposure and infant growth also found inconsistent results (Tesfamariam et al., 2020). With the variation in the detected types and concentrations of mycotoxins and confounding factors adjusted in previous literature, It’s understandable that only certain studies found.

Despite the lack of apparent association between OTA exposure and newborn and infant anthropometry in the present study, the high prevalence of OTA exposure can potentially have severe adverse consequences. After its absorption from the gastrointestinal tract, OTA binds mainly to albumin with high affinity, resulting in its long half-life (Kőszegi & Poór, 2016). Based on previous studies, The OTA mechanism is understood to be carcinogenic, hepatotoxic, immunotoxic, neurotoxic and teratogenic (el Khoury & Atoui, 2010; Ringot et al., 2006). In humans, OTA exposure has been associated with adverse health effects such as Tunisian Nephropathy (Hassen et al., 2004), esophageal and gastric tumors (Cui et al., 2010; Liu et al., 2015), as well as testicular cancer (Schwartz, 2002).

Given the risks associated with mycotoxins exposure in both the current population and similar demographics, Matumba *et al*. (2021) put forward an extensive strategy for the prevention and management of mycotoxin contamination in grains, specifically tailored for LMICs This framework comprises five key recommendations, which include: i) Enhancing the strength and health of plants; ii) Decreasing the population of toxigenic fungi during plant growth and storage; iii) Swiftly reducing the moisture content of grains and avoiding rehydration; iv) Protecting the outer structure of seeds/grains; and v) Thoroughly cleaning and removing components at high risk of mycotoxin contamination. (Matumba et al., 2021).

The internal method validation showed a good performance of mycotoxin detection as suggested by parameters including the LOD, LLOQ, apparent recovery, RSDr and RSDR parameters. These findings are also supported by a previous validation conducted in the same lab (Vidal et al., 2021). To our understanding, this marks the first use of VAMS for mycotoxin analysis in the whole blood of pregnant women within a LMIC setting. Considering the benefits provided by VAMS and the resilient method established previously, VAMS sampling can serve as an alternative method to venous sampling for conducting quantitative screening of mycotoxin exposure (Vidal et al., 2021). In addition, considering the toxicokinetic profiles of the detected mycotoxins, this microsampling technique will additionally emphasize the impact of mycotoxin exposure on human health, facilitating associations to be interpreted with adverse health effects.

### Strengths and limitations

Some of the strengths of the present study are the timely and robust assessment of study outcomes including determination of GA using ultrasonography and the assessment of birth outcomes within 12 hours of birth. Despite the prospective design of the study, mycotoxin exposure was determined during the third trimester of pregnancy leading to a narrow time gap between exposure and outcomes assessment. Direct determination of mycotoxin exposure using biomarkers is superior to the assessment in foodstuffs used in other studies (De Boevre et al., 2013). The VAMS procedure used for sampling in the present study facilitated sample collection, wherein a delicate finger or heel incision (for infants less than 10 kilos) is required without participants travelling long distances to clinical services, and in addition, the small sample volumes collected is favorable for anemic participants and newborns. VAMS provided additional advantages concerning sample handling, storage, and transportation. The procedure has also been shown to be robust and valid for assessing mycotoxin exposure (Vidal et al., 2021) since the UPLC-MS/MS analytical method was previously reported to be highly sensitive and specific (Tesfamariam et al., 2022). However, only a limited number of biomarkers have undergone validation for assessing mycotoxin exposure, specifically AFM1, AFB1-N7-guanine, and DON-glucuronides in urine, as well as the adduct of AFB1 with albumin and AFB1-lysine in human plasma (Vidal et al., 2018). In our study, exposure to AFB1-lysine was not assessed due to the current unavailability of the commercial standard, for this reason only the free mycotoxins AFB1, AFB2 AFG1 AFG2 and AFM1 were measured. Therefore, results obtained using non-validated biomarkers (such as whole blood OTA) should be approached with caution. Furthermore, there are alternative matrices to blood which offer interesting advantages. Even though present at a lower concentration in urine than in blood, OTA in urine was demonstrated in some studies to be a better indicator of exposure to OTA (Castegnaro et al., 2006; Gilbert et al., 2001; Pfohl-Leszkowicz A et al., 2006). In a study employing both serum and urinary biomarkers of OTA exposure, a more robust correlation was observed between dietary OTA intake and urinary OTA levels compared to serum OTA levels (Gilbert et al., 2001). Lastly, mycotoxin exposure data from only a single time point during pregnancy was considered. Future studies, using repeated mycotoxins measurements, will provide an insight into the effects of mycotoxins and their physicochemical properties in relation to the timing of exposure.

## Conclusion

Although the present study did not yield any evidence of associations between OTA and birth outcomes or infant growth, we found evidence for a significant presence of OTA exposure among pregnant participants. Public health policies and nutrition-sensitive interventions need to prioritize the reduction of mycotoxin exposure in rural Burkina Faso.

## Data availability request

Due to the sensitive nature of the data, access to the data will be facilitated through a data-sharing agreement. For inquiries, please reach out to. Additionally, supporting study documents such as the study protocol and questionnaires can be accessed publicly on the study’s website: https://misame3.ugent.be (accessed on 07 December 2023).

## Statements and Declarations

### Author contributions

Author Contributions: Author Conceptualization: Y.B.-M., A.A., T.D.-C., S.D.S.,CL and M.D.B.; Methodology: Y.B-M., J-E-H., S.D.S. and M.D.B.; Software: Y.B.-M., A.A., and T.D.-C.; Validation: Y.B-M., J-E-H. and M.D.B.; Formal analysis: Y.B-M and A.A.; Investigation: Y.B.-M., T.D.-C, J.E-H., L.O. and M.D.B.; Resources: Y.B.-M., T.D.-C., L.O., L.C.T., S.D.S, C.L. and M.D.B.; Data Curation: Y.B.-M., A.A., T.D.-C. and L.O.; Writing—Original Draft: Y.B.-M. and A,A.; Writing—Review and Editing: Y.B.-M., A.A., T.D.-C., J.E-H., L.O., L.C.T., S.D.S., C.L and M.D.B.; Supervision: T.D.-C., S.D.S, C.L., and M.D.B.; Project Administration: Y.B.-M., T.D.-C., L.O., C.L., S.D.S. and M.D.B.; Funding Acquisition: T.D.-C., C.L. and M.D.B.

### Ethical considerations

The study protocol received approval from the Ethical Committee of Ghent University Hospital in Belgium (B670201734334) and the Ethical Committee of Institut de Recherche en Sciences de la Santé in Burkina Faso (50-2020/CEIRES). All participants provided written informed consent prior to their involvement in the study.

### Consent to participate

All participants in the study provided informed consent.

### Consent for publication

All the authors also agreed on the publication of this article.

## Abbreviations

AFs: aflatoxins
AFG1: aflatoxin G1
AFM1: aflatoxin M1
AIC: Akaike Information Criterion
BEP: balanced energy-protein
BIC: Bayesian Information Criterion
BioSpé: Biospecimen
CPA: cyclopiazonic acid
DON: deoxynivalenol
FBs: fumonisins
FB1: fumonisin B1
GA: gestational age
IARC: International Agency for Research on Cancer
IFA: iron-folic acid
LAZ: length-for-age z-score
LBW: low birth weight
LMICs: low-and-middle income countries
LOD: limit of detection
LLOQ: lower limit of quantification
MISAME: MIcronutriments pour la SAnté de la Mère et de l’Enfant
MUAC: mid-upper arm circumference
OTA: ochratoxin A
PTB: preterm birth
RSDr: Repeatability
RSDR: reproducibility
SGA: small-for-gestational age
UPLC-MS/MS: ultra-performance liquid chromatography-tandem mass spectrometry
VAMS: volumetric absorptive microsampling
WAZ: weight-for-age z score
WLZ: weight-for-length z score.

